# An Initial Genetic Correlation Analysis of Externalizing Behavior and Neuroimaging Phenotypes in the ABCD Cohort

**DOI:** 10.64898/2026.07.13.26358013

**Authors:** Mengman Wei, Qian Peng

## Abstract

Adolescent externalizing behavior is a major risk factor for later substance use and other psychiatric outcomes. Understanding its genetic architecture and its relationships with brain imaging phenotypes requires scalable genome-wide methods that can be applied to youth cohorts. Using data from the Adolescent Brain Cognitive Development (ABCD) Study^®^, we implemented a pipeline for conducting genome-wide association studies (GWAS) of longitudinal externalizing traits and multimodal imaging-derived phenotypes (IDPs). We performed quality-controlled genotype processing and constructed harmonized phenotype and covariate datasets. GWAS analyses were conducted using REGENIE in a two-step framework. In Step 1, ridge regression prediction models were trained using linkage disequilibrium (LD)-pruned variants. In Step 2, genome-wide association testing was performed for each phenotype. The analyses included three externalizing phenotypes—baseline, longitudinal mean, and longitudinal slope—and approximately 200 IDPs measured at baseline or summarized using their longitudinal means and slopes. We additionally constructed a custom LD reference panel using unrelated individuals and calculated LD scores using LD Score Regression software (LDSC). Genome-wide genetic correlations between externalizing traits and imaging phenotypes were subsequently estimated using cross-trait LD Score Regression.

This exploratory study systematically evaluated genome-wide genetic correlations between regional cortical morphology and externalizing phenotypes during adolescence. Although several associations reached nominal statistical significance, none remained significant after correction for multiple comparisons. These results should not be interpreted as evidence for the absence of shared genetic architecture. Instead, the precision of the genetic-correlation estimates was limited by the available imaging GWAS sample size, uncertainty in SNP-based heritability estimates, and the large number of regional comparisons. Larger imaging-genetics samples and independent replication studies will be required to determine whether modest or regionally specific genetic correlations exist.

## Introduction

Externalizing behavior refers to a broad set of outwardly directed behavioral tendencies, including impulsivity, rule-breaking, aggression, and related forms of behavioral dysregulation [1, 2]. These traits are particularly important during adolescence because elevated externalizing behavior is associated with later mental health problems, increased risk of substance use, and broader developmental difficulties. At the same time, adolescence is a period of rapid and heterogeneous brain development, making it a critical developmental window for studying how behavioral risk is related to neurobiological variation [3, 4, 5].

The Adolescent Brain Cognitive Development (ABCD) Study is well suited to addressing this question because it follows a large cohort of children from approximately 9–10 years of age into adolescence and includes comprehensive behavioral assessments, neuroimaging data, environmental measures, and genome-wide genotype data [6, 7, 8, 9, 10]. The ABCD Study was designed as a large, longitudinal, multisite study in the United States. Its standardized neuroimaging protocol supports harmonized measurements of brain structure and function across study sites, scanner platforms, and manufacturers. These features make the ABCD Study a valuable resource for investigating the genetic and neurobiological contributions to behavioral development during adolescence.

A central question is whether externalizing behavior and brain imaging-derived phenotypes (IDPs) share common genetic influences. Genome-wide association studies (GWAS) can be used to estimate variant-level associations with externalizing behavior and individual imaging phenotypes [11]. Cross-trait linkage disequilibrium (LD) Score Regression can subsequently estimate the genome-wide genetic correlation between two sets of GWAS summary statistics [12]. This method provides a scalable approach for testing whether externalizing behavior and specific IDPs show evidence of shared polygenic architecture. It is particularly useful when many IDPs must be evaluated and when the objective is to identify brain-related traits that are genetically correlated with externalizing liability, rather than traits that are only phenotypically associated with externalizing behavior.

In this study, we describe a complete analytical workflow for estimating genetic correlations between externalizing behavior and IDPs in the ABCD Study. The workflow has four primary goals. First, it prepares harmonized phenotype and covariate files for genome-wide genetic association analyses. Second, it performs scalable two-step GWAS using REGENIE, a whole-genome regression framework designed for large-scale genetic association studies [13]. Third, it constructs a study-specific LD Score reference panel using unrelated participants from the same pooled genotype resource. Fourth, it harmonizes the resulting GWAS summary statistics and estimates genome-wide genetic correlations between externalizing behavior and brain imaging phenotypes. Together, this workflow provides a reproducible framework for identifying IDPs that share genetic architecture with externalizing behavior and may help prioritize neurobiological systems relevant to behavioral risk during adolescence.

## Methods

### Study cohort

Data were obtained from the Adolescent Brain Cognitive Development (ABCD) Study [6, 7, 8, 9, 10], a large prospective longitudinal study of child and adolescent health and development in the United States. The ABCD Study recruited approximately 11,880 children who were 9–10 years old at baseline from multiple research sites across the United States. The present study used ABCD Curated Data Release 5.1, which includes longitudinal behavioral assessments, structural magnetic resonance imaging measures, demographic and technical information, and genome-wide genotype data.

The genotype dataset used in the present analyses contained 11,666 participants after genotype quality-control procedures. Participants were eligible for a phenotype-specific genome-wide association analysis when they had genotype data, the phenotype being analyzed, and the covariates required for that analysis. Because behavioral, imaging, and covariate availability differed across participants and assessment waves, the final analytic sample varied across phenotypes.

For the externalizing analyses, 11,664 participants were present in the intersection of the genotype, covariate, and externalizing phenotype datasets before phenotype-specific missingness was considered. Three externalizing phenotypes were analyzed: externalizing behavior at baseline, the longitudinal mean of externalizing behavior across available assessments, and the longitudinal slope of externalizing behavior over time. The final REGENIE genome-wide association analyses included 11,088 participants for baseline externalizing behavior, 11,093 participants for the longitudinal mean, and 10,783 participants for the longitudinal slope.

For the cortical imaging analyses, 11,533 participants were present in the intersection of the genotype, cortical imaging, covariate, and intracranial-volume datasets before phenotype-specific missingness was considered. The final imaging analyses included 198 cortical phenotypes representing cortical surface area and cortical thickness at baseline, their longitudinal means, and their longitudinal slopes. Although the broader imaging phenotype table contained 216 columns during the initial REGENIE Step 1 model fitting, 198 distinct imaging phenotypes were carried forward to the final Step 2 genome-wide association analyses and subsequent genetic-correlation analyses.

Intracranial volume was included as an additional covariate in the cortical surface-area analyses. After accounting for phenotype-specific availability, the REGENIE-reported sample sizes for the 198 imaging GWAS ranged from 7,995 to 11,093 participants, with a median sample size of 11,093. Exact sample sizes for individual imaging phenotypes are reported in the Supplementary Materials.

The combined set of participants included in at least one of the analyzed externalizing or imaging phenotypes comprised 11,093 individuals. Their mean baseline age was 9.48 years (SD = 0.51; range, 8–11 years). Age, sex, study site, and genetic principal components were included as covariates as specified for each genome-wide association analysis. Imaging analyses additionally incorporated prespecified imaging-acquisition and quality-control covariates, including head-motion and scanner-related variables. Intracranial volume was additionally included for cortical surface-area phenotypes.

All analyses were secondary analyses of de-identified ABCD Study data and were conducted in accordance with the ABCD data-use agreement and applicable institutional and data-access requirements.

### Genotype data and quality control

Genome-wide imputed genotype data were processed using PLINK version 1.90b7.2 [14]. The initial imputed dataset contained 280,985,564 variants and 11,666 participants. Genotype data were stored in PLINK binary format.

Participant-level quality control excluded individuals with more than 5% missing genotype data. No participants were removed by this criterion. Variant-level quality control retained variants with a minor allele frequency (MAF) of at least 0.01, a genotype missingness rate of no more than 0.05, and a Hardy–Weinberg equilibrium mid-*P* value of at least 1 × 10^−6^. After these filters, the genotype dataset contained 8,118,068 variants and 11,666 participants. This quality-controlled, unpruned dataset was used as the primary genotype dataset for the genome-wide association analyses.

A reduced set of approximately independent variants was created for REGENIE Step 1 whole-genome prediction [15]. Variants used for Step 1 were required to have an MAF of at least 0.01, genotype missingness of no more than 0.01, and a Hardy–Weinberg equilibrium mid-*P* value of at least 1×10^−6^. Linkage disequilibrium (LD) pruning was performed using a 1,000-kb window and a step size of 50 variants. An initial *r*^2^ threshold of 0.20 was used for the imaging Step 1 model and was reduced to 0.10 when necessary to obtain no more than one million variants. The final imaging Step 1 model included 605,775 LD-pruned variants.

For the externalizing Step 1 model, variants in predefined long-range LD regions were excluded before pruning because extended LD can affect the construction of approximately independent variant sets [16]. These regions included the major histocompatibility complex on chromosome 6 and additional long-range LD regions on chromosomes 8, 11, and 17. LD pruning was then performed using the same 1,000-kb window and 50-variant step size, with progressively more stringent *r*^2^ thresholds when required to keep the Step 1 training set below one million variants.

The LD-pruned variants were used only to fit the REGENIE Step 1 prediction models. In REGENIE Step 2, genome-wide association testing was performed using the full set of 8,118,068 quality-controlled imputed variants rather than the LD-pruned subset. Variants with a minor allele count (MAC) below 20 were excluded from the Step 2 association tests.

### Phenotype construction

#### Externalizing phenotypes

Externalizing behavior was measured using the Child Behavior Checklist externalizing-problems T-score [17], represented in ABCD by the variable `cbcl_scr_syn_external_t`. Measurements were obtained from the baseline, 1-year, 2-year, 3-year, and 4-year assessments when available.

Three subject-level externalizing phenotypes were constructed. `EXT_baseline` was the externalizing T-score recorded at the baseline assessment. `EXT_mean` was the arithmetic mean of all available, nonmissing externalizing T-scores across the five included assessment waves. Participants with at least one valid externalizing measurement could contribute to the longitudinal mean.

`EXT_slope` represented the participant-specific yearly change in externalizing T-score. Assessment waves were coded as 0, 1, 2, 3, and 4 years from baseline. For each participant with at least two valid measurements obtained at different assessment times, an ordinary least-squares regression of externalizing T-score on years since baseline was fitted. The estimated regression coefficient for time was retained as the longitudinal slope. A positive slope indicated an increase in externalizing T-score over time, whereas a negative slope indicated a decrease.

Scores that could not be converted to numeric values were treated as missing. Phenotype construction was performed without adjustment for covariates; covariates were included later in the genome-wide association analyses. The resulting subject-level phenotype table contained the participant identifier and the variables `EXT_baseline`, `EXT_mean`, and `EXT_slope`. During preparation for REGENIE, participants were restricted to those present in the required genotype and covariate datasets, and family and individual identifier columns were placed at the beginning of the phenotype file. The externalizing slope used nominal visit time, coded as 0, 1, 2, 3, and 4 years from baseline, rather than each child’s exact age. For participant *i*, the slope can be written as

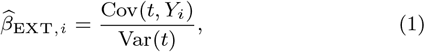

where *Y*_*i*_ denotes the participant’s observed externalizing T-scores and *t* denotes nominal years since baseline.

#### Cortical imaging phenotypes

Cortical regions were defined using the Desikan–Killiany cortical parcellation, which divides each cerebral hemisphere into 34 gyral regions [18]. Homologous left- and right-hemisphere measurements were combined to produce 34 bilateral cortical regions. For each region, cortical surface area and cortical thickness were examined, resulting in 68 regional morphometric measures. A global surface-area summary and an area-weighted global cortical-thickness measure were also constructed, giving 70 underlying imaging measures. For each measure, baseline, longitudinal-mean, and longitudinal-slope phenotypes were generated, resulting in 210 unique longitudinal imaging phenotypes.

Longitudinal cortical imaging phenotypes were constructed from the ABCD structural MRI tables, including the cortical surface-area and cortical-thickness tables. Surface-area measures were obtained from `mri_y_smr_area_dsk.csv`, and cortical-thickness measures were obtained from `mri_y_smr_thk_dsk.csv`. Records from the two tables were matched by participant and assessment visit. Age in years at each imaging visit was then added from a separate participant-by-visit age table.

Only cortical regions with both left- and right-hemisphere surface-area and thickness measurements were retained. For each region and visit, bilateral surface area was calculated by summing the left- and right-hemisphere values. Bilateral cortical thickness was calculated as the mean of the left- and right-hemisphere values. Values that could not be converted to numeric form were treated as missing.

For each regional surface-area and cortical-thickness measure, three subject-level longitudinal summaries were created. The baseline phenotype, identified by the suffix `_baseline`, was the value recorded at the ABCD baseline imaging visit. The longitudinal mean, identified by `_mean`, was the arithmetic mean of all available, nonmissing imaging measurements for that participant. The longitudinal slope, identified by `_slope`, was estimated by fitting an ordinary least-squares regression of the imaging measure on the participant’s age in years at each imaging visit. At least two visits with valid age and imaging measurements were required to estimate a slope. A positive slope indicated an increase in the imaging measure with age, whereas a negative slope indicated a decrease.

The construction script initially generated baseline, mean, and slope columns for the available cortical measures. Of the 210 unique longitudinal imaging phenotypes constructed, 198 distinct cortical imaging phenotypes were included in the final REGENIE Step 2 analyses. Regional surface-area phenotypes were identified by `_SA_`, and cortical-thickness phenotypes were identified by `_TH_`. During preparation for REGENIE, the imaging table was restricted to participants with the required genotype, phenotype, covariate, and intracranial-volume information. Family and individual identifier columns were then added as the first two columns of the final imaging phenotype file.

The imaging slope used each participant’s actual age in years at each imaging visit, rather than nominal visit time. For participant *i*, the slope can be written as

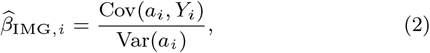

where *Y*_*i*_ denotes the participant’s observed imaging measurements and *a*_*i*_ denotes the participant’s age in years at the corresponding imaging visits. Thus, the externalizing and imaging slopes were constructed differently: the externalizing slope used nominal years since baseline, whereas the imaging slope used each participant’s actual age at each imaging visit.

#### Covariates

Association models adjusted for demographic, technical, and genetic covariates that could influence the behavioral or imaging phenotypes. The primary covariates were age, sex, study site, scanner manufacturer, scanner device serial number, head motion, and the first 20 genetic principal components.

Study site, sex, scanner manufacturer, and scanner device serial number were treated as categorical covariates. Age, head motion, and genetic principal components were treated as continuous covariates. Estimated total intracranial volume was included as an additional continuous covariate for cortical surface-area phenotypes. Cortical-thickness models did not include estimated total intracranial volume. Before association testing, the covariate files were checked for malformed or merged column names, including incorrectly joined principal-component headers. Covariate columns expected to be numeric were converted and validated as numeric variables. Categorical and continuous covariates were then supplied separately to REGENIE.

#### Sample harmonization

Participant identifiers were harmonized across genotype, phenotype, imaging, and covariate files before association analysis.

For the externalizing GWAS, participants were required to be present in the following datasets:

1. the genotype dataset;
2. the cleaned covariate dataset; and
3. the externalizing phenotype dataset.

For the imaging GWAS, participants were required to be present in the following datasets:

1. the genotype dataset;
2. the cleaned imaging covariate dataset;
3. the imaging phenotype dataset; and
4. the intracranial-volume dataset, when required.

Two PLINK-compatible participant-inclusion files were created: `keep.ext` for the externalizing analyses and `keep.img` for the imaging analyses. Missing phenotype values were handled separately for each phenotype, resulting in phenotype-specific analytic sample sizes.

#### Genome-wide association analysis

Genome-wide association analyses were performed using REGENIE [15], which uses a two-step whole-genome regression framework. All externalizing and imaging outcomes were analyzed as quantitative phenotypes.

##### REGENIE Step 1: whole-genome prediction

In Step 1, REGENIE fitted ridge-regression prediction models using the quality-controlled, LD-pruned genotype dataset. Leave-one-chromosome-out predictions generated in Step 1 were saved for use in Step 2. Separate Step 1 analyses were conducted for the three externalizing phenotypes and for the complete set of cortical imaging phenotypes.

Low-memory mode was enabled to reduce computational memory requirements. If the initial pruned genotype set contained more than one million variants, more stringent LD pruning was applied before fitting the Step 1 models.

##### REGENIE Step 2: genome-wide association testing

In Step 2, genetic variants from the complete quality-controlled imputed genotype dataset were tested for association with each phenotype. The Step 1 leave-one-chromosome-out predictions were included to account for polygenic background and relatedness.

Association testing used the following REGENIE options:

~~~
--bsize 400
--minMAC 20
--gz
~~~

Thus, variants were tested only when the minor allele count was at least 20. Results were written as compressed REGENIE summary-statistic files.

The three externalizing phenotypes were analyzed separately. The 198 imaging phenotypes were distributed across independent SLURM array tasks, with one imaging phenotype analyzed per task. Surface-area phenotypes used the covariate file containing estimated total intracranial volume, whereas cortical-thickness phenotypes used the base imaging covariate file.

#### Construction of the linkage disequilibrium reference panel

A study-specific LD reference panel was constructed from the same pooled genotype resource used for the genome-wide association analyses.

An unrelated subset of participants was selected before LD estimation. Related individuals were removed using the KING-robust kinship estimator implemented in PLINK 2 [19], with a kinship-coefficient cutoff of 0.0884. This threshold excludes pairs of participants related at approximately the second-degree level or closer [20].

The unrelated genotype panel was divided into chromosome-specific PLINK datasets for autosomes 1–22. Chromosome-specific LD Scores were then calculated using LD Score Regression with a 1,000-kb LD window [12]:

`ldsc.py --l2 --ld-wind-kb 1000`

The resulting chromosome-specific files used the prefix `pooled.custom`. The custom LD Scores were used as both the reference LD Scores and the regression weights in the primary genetic-correlation analyses.

#### Harmonization of GWAS summary statistics

Externalizing and imaging GWAS summary statistics were converted to a common LD Score Regression format using a direct harmonization procedure. Variant identifiers were assigned using chromosome-position-allele keys of the form

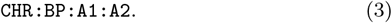

Each key was mapped to an rsID, and variants were then restricted to those represented in the custom LD reference panel.

The harmonization procedure performed the following steps:

1. standardized the chromosome, position, allele, effect-size, standard-error, *P*-value, and sample-size fields;
2. calculated the signed *Z* statistic as

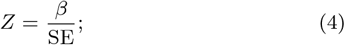
3. mapped chromosome-position-allele combinations to rsIDs;
4. removed variants absent from the reference panel;
5. removed invalid, non-biallelic, or strand-ambiguous variants;
6. aligned effect and non-effect alleles to the reference panel;
7. reversed the sign of the *Z* statistic when the allele orientation was reversed; and
8. removed duplicate variants and records with invalid *Z* statistics or sample sizes.

The resulting files contained the following columns:

`SNP N Z A1 A2` SNP-level sample sizes from the REGENIE results were retained. Each harmonized file was required to contain at least 200,000 variants before it was accepted for downstream analysis. Preparation quality-control files recorded the numbers of input variants, retained variants, excluded variants, allele mismatches, duplicate variants, and allele-orientation changes.

#### SNP-heritability estimation

LD Score Regression was used to estimate SNP heritability for each externalizing and cortical imaging phenotype. For each phenotype, the following quantities were recorded:

- the SNP-heritability estimate;
- its standard error;
- the SNP-heritability *Z* statistic;
- the mean GWAS chi-square statistic;
- the genomic-control inflation factor;
- the LD Score Regression intercept; and
- the number of regression variants.

These values were used to evaluate the reliability of the genetic-correlation estimates. The primary analysis retained technically estimable genetic correlations without imposing a strict heritability-*Z* threshold. A sensitivity analysis was subsequently restricted to comparisons in which both phenotypes had SNP-heritability *Z* statistics of at least 4. This sensitivity analysis was used to determine whether the overall conclusions were similar when attention was restricted to phenotypes with more precisely estimated SNP heritability.

#### Genetic-correlation analysis

Cross-trait genetic correlations were estimated using LD Score Regression [21]. Genetic correlation measures the extent to which common-variant effects are shared between two traits and was defined as

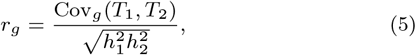

where Cov_*g*_(*T*_1_, *T*_2_) is the estimated genetic covariance between traits *T*_1_ and *T*_2_, and 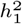 and 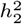 are their SNP-heritability estimates.

Each of the three externalizing phenotypes was compared with each of the 198 cortical imaging phenotypes, producing

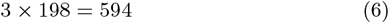

planned genetic-correlation analyses.

For each phenotype pair, LD Score Regression was run using the harmonized summary-statistic files and the study-specific LD reference and weight files:

~~~
ldsc.py \
--rg imaging.sumstats.gz,externalizing.sumstats.gz \
--ref-ld-chr pooled.custom. \
--w-ld-chr pooled.custom. \
--out output_prefixv
~~~

For each comparison, the genetic-correlation estimate, standard error, *Z* statistic, and *P* value were extracted. A comparison was considered estimable when these statistics were finite and could be parsed from the LD Score Regression output. Undefined estimates were retained in the analysis records but were classified as non-estimable rather than nonsignificant. Genetic correlation represents shared common-variant architecture and does not establish a causal relationship between cortical morphology and externalizing behavior [22].

#### Confidence intervals and multiple-testing correction

A two-sided 95% confidence interval (CI) was calculated for each estimable genetic correlation:

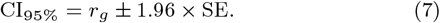

Nominal statistical significance was defined as *P* < 0.05. Because hundreds of correlated phenotype pairs were tested, nominal *P* values were treated as exploratory.

The primary multiple-testing correction used the Benjamini– Hochberg false-discovery-rate procedure [23] across all estimable genetic-correlation *P* values. Associations with a Benjamini– Hochberg-adjusted *P* < 0.05 were considered statistically significant after global false-discovery-rate correction.

As secondary descriptive analyses, adjusted *P* values were also calculated separately within each externalizing phenotype. These within-phenotype corrections were not used as substitutes for the primary global correction. For the heritability-quality sensitivity subset, false-discovery-rate correction was recalculated among comparisons for which both phenotypes had SNP-heritability *Z* statistics of at least 4.

#### Equivalence analysis

Failure to reject the conventional null hypothesis *r*_*g*_ = 0 does not demonstrate that a genetic correlation is absent or sufficiently small. Therefore, equivalence testing was conducted to identify comparisons for which the data supported genetic correlations smaller than a prespecified magnitude [24].

The equivalence region was defined as

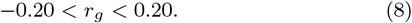

Thus, the lower and upper equivalence bounds were set to −0.20 and 0.20, respectively. These bounds represented the largest absolute genetic correlation considered substantively negligible for the purposes of this study.

Equivalence was evaluated using the two one-sided tests procedure [24, 25]. At a one-sided significance level of *α* = 0.05, a genetic correlation was classified as statistically equivalent when its two-sided 90% CI was fully contained within the prespecified equivalence region:

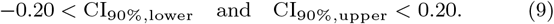

The 90% CI was calculated as

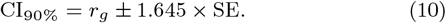

Comparisons that were neither statistically different from zero nor statistically equivalent were classified as inconclusive. This category indicates that the estimate was not sufficiently precise to support either a nonzero genetic correlation or equivalence within the prespecified bounds [24].

The equivalence margin of |*r*_*g*_| = 0.20 was prespecified before examination of the genetic-correlation results. It was treated as a threshold for the smallest genetic-correlation magnitude considered substantively important, rather than as a universal benchmark for genetic correlations.

#### Sensitivity and quality-control analyses

Several additional analyses were conducted to evaluate the reliability of the results.

First, genetic correlations were summarized separately according TO

- cortical thickness and cortical surface area;
- baseline, longitudinal-mean, and longitudinal-slope phenotypes; and
- each externalizing phenotype.

Second, the analysis was repeated within the subset in which both traits had SNP-heritability *Z* statistics of at least 4. Third, non-estimable comparisons were examined separately to determine whether they were concentrated among imaging phenotypes with weak or unstable SNP-heritability estimates. Fourth, the distributions of standard errors and confidence-interval widths were examined to distinguish evidence of small effects from insufficient statistical precision.

Unless accompanied by a prespecified formal statistical test, subgroup comparisons, distributional summaries, and visualizations were interpreted descriptively.

## Results

### SNP heritability

All three externalizing phenotypes had positive and statistically informative SNP-heritability estimates. Estimated SNP heritability was 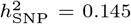 for baseline externalizing behavior (SE = 0.034, *Z* = 4.32), 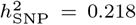 for mean externalizing behavior (SE = 0.032, *Z* = 6.81), and 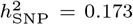 for longitudinal externalizing slope (SE = 0.038, *Z* = 4.55).

SNP-heritability statistics were successfully obtained for 196 of the 198 imaging phenotypes. Of these, 167 had heritability *Z* statistics of at least 2, and 89 had heritability *Z* statistics of at least 4. Cortical-thickness phenotypes generally had more precisely estimated SNP heritability than baseline and mean surface-area phenotypes. Median heritability *Z* statistics were 4.46, 4.86, and 4.68 for baseline, mean, and slope cortical-thickness phenotypes, respectively. The corresponding median values for surface-area phenotypes were 2.66, 2.62, and 4.45. However, surface-area slopes showed substantial variability and included several negative heritability estimates.

### Genetic correlations between externalizing behavior and cortical imaging phenotypes

Among the 576 estimable genetic correlations, 42 (7.3%) had unadjusted *P* < 0.05. However, no individual association remained statistically significant after global Benjamini–Hochberg false-discovery-rate correction. No association survived the secondary false-discovery-rate correction performed separately within each externalizing phenotype.

The strongest nominal association was observed between mean externalizing behavior and longitudinal change in entorhinal cortical thickness:

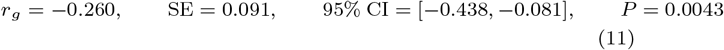

The negative estimate indicates that common variants associated with higher mean externalizing behavior tended to be associated with a more negative, or less positive, age-related change in entorhinal cortical thickness. This result was nominal and did not remain significant after multiple-testing correction.

Other highly ranked nominal results included a negative genetic correlation between mean insular cortical thickness and externalizing slope (*r*_*g*_ = −0.404, *P* = 0.0045), as well as positive genetic correlations between baseline superior frontal cortical thickness and both baseline externalizing behavior (*r*_*g*_ = 0.367, *P* = 0.0045) and mean externalizing behavior (*r*_*g*_ = 0.316, *P* = 0.0049). These associations also did not survive false-discovery-rate correction.

Nominal associations were observed more frequently for cortical-thickness phenotypes than for surface-area phenotypes, with 32 and 10 nominal associations, respectively. Eighteen nominal associations involved imaging-slope phenotypes, compared with 11 involving baseline imaging phenotypes and 13 involving longitudinal imaging means. By externalizing definition, 19 nominal associations involved baseline externalizing behavior, 17 involved mean externalizing behavior, and 6 involved externalizing slope. These subgroup patterns were descriptive and were not tested as formal differences between phenotype groups.

Across all imaging phenotypes, the distributions of genetic-correlation estimates were centered close to zero for each externalizing outcome. The median genetic correlations were −0.008 for baseline externalizing behavior, −0.024 for mean externalizing behavior, and −0.017 for externalizing slope. Nevertheless, many estimates had wide confidence intervals, limiting the ability to distinguish small genetic correlations from imprecisely estimated larger effects.

### Heritability-based sensitivity analysis

A sensitivity analysis was performed among comparisons for which both the imaging and externalizing phenotypes had SNP-heritability *Z* statistics of at least 4. This criterion retained 267 of the 576 estimable correlations. Twenty-six correlations had nominal *P* < 0.05 in this restricted dataset, but none remained significant after false-discovery-rate correction within the sensitivity subset.

The strongest association in the sensitivity analysis remained the genetic correlation between entorhinal cortical-thickness slope and mean externalizing behavior. Its false-discovery-rate-adjusted *q* value within the sensitivity subset was 0.212. Therefore, restricting the analysis to phenotypes with more precisely estimated SNP heritability did not materially alter the primary conclusion.

### Equivalence analysis

Equivalence testing was used to determine whether the data were sufficiently precise to support genetic correlations within the prespecified negligible-effect interval

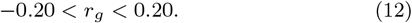

Twenty-two of the 576 estimable correlations (3.8%) had 90% confidence intervals fully contained within these bounds and were therefore statistically equivalent to an absolute genetic correlation smaller than 0.20. Twelve of these involved surface-area slopes, while the remaining 10 involved cortical-thickness phenotypes.

When the conventional and equivalence tests were considered together, 42 correlations showed nominal evidence of being different from zero, 22 supported equivalence within the prespecified bounds, and 512 (88.9%) were inconclusive. Thus, most nonsignificant findings did not provide sufficient evidence that the corresponding genetic correlations were absent or smaller than 0.20.

## Discussion

This study systematically evaluated common-variant genetic correlations between longitudinal externalizing behavior and regional cortical surface-area and cortical-thickness phenotypes in the ABCD cohort. Although 42 imaging–externalizing comparisons reached nominal significance, none remained significant after correction for the full set of comparisons. The strongest exploratory result was a negative genetic correlation between mean externalizing behavior and age-related change in entorhinal cortical thickness. However, its global false-discovery-rate-adjusted *q* value was 0.403, and it should therefore be considered a hypothesis-generating observation rather than evidence for a reproducible regional association.

Importantly, the absence of associations surviving multiple-testing correction should not be interpreted as evidence that externalizing behavior and cortical morphology have no shared genetic architecture. Equivalence testing supported an absolute genetic correlation below 0.20 for only 22 of the 576 estimable comparisons. Most comparisons were neither statistically different from zero nor statistically equivalent within the prespecified bounds. The dominant result was therefore statistical uncertainty, rather than affirmative evidence of no relationship. This distinction is important because failure to reject *r*_*g*_ = 0 does not demonstrate that the true correlation is zero or negligibly small.

Nominal results were more frequent among cortical-thickness phenotypes than among surface-area phenotypes. Several of the most highly ranked associations involved superior frontal, entorhinal, insular, postcentral, and pars opercularis cortical-thickness measures. Nevertheless, the associations did not form a sufficiently consistent pattern to support strong regional interpretation. Some regions also showed different directions across externalizing definitions or across baseline and longitudinal imaging summaries. Given the large number of correlated comparisons and the absence of corrected significance, interpreting individual nominal regions as specific neurobiological mechanisms would be premature.

Differences in statistical precision may partly explain the observed pattern. Cortical-thickness phenotypes generally had higher SNP-heritability *Z* statistics than baseline and mean surface-area phenotypes. Because genetic correlation is calculated from genetic covariance relative to the SNP heritabilities of both traits, uncertainty in either heritability estimate can produce imprecise or unstable genetic-correlation estimates. The sensitivity analysis restricted to traits with heritability *Z ≥* 4 produced the same overall conclusion, indicating that the lack of corrected associations was not explained solely by the phenotypes with the weakest heritability estimates. Nevertheless, even within this restricted subset, the confidence intervals remained too wide to identify modest correlations reliably.

Longitudinal slope phenotypes presented an additional challenge. All non-estimable genetic correlations involved imaging slopes, and several of these imaging traits had negative or unavailable SNP-heritability estimates. A longitudinal slope is estimated from differences across repeated measurements and is therefore especially sensitive to measurement error, the number and timing of available visits, missing assessments, and individual variation in follow-up duration. The externalizing slopes were calculated using nominal visit times, whereas imaging slopes were calculated using each participant’s actual age. Although both approaches estimate longitudinal change, this difference should be considered when comparing their genetic architectures.

The findings also need to be interpreted in relation to the developmental period represented by ABCD. Brain structure and externalizing behavior both change considerably during late childhood and adolescence. A genetic correlation involving a baseline measure may therefore represent a different developmental process from a correlation involving a longitudinal mean or slope. Combining left- and right-hemisphere measurements into bilateral phenotypes improved the manageability of the analysis but may also have obscured hemisphere-specific genetic associations.

This study has several strengths. It used a large, deeply phenotyped longitudinal youth cohort and evaluated baseline levels, longitudinal means, and developmental slopes rather than relying on a single behavioral or imaging assessment. Genotype, phenotype, and covariate data were processed through a consistent GWAS workflow, and genetic correlations were estimated using harmonized summary statistics and a study-specific LD reference. Non-estimable results were distinguished from nonsignificant results, multiple testing was controlled globally, and equivalence testing was used to distinguish evidence for small effects from a simple lack of statistical significance.

Several limitations should also be acknowledged. First, although ABCD is large for a longitudinal neuroimaging study, the imaging GWAS sample sizes remain small relative to those typically required for precise genetic-correlation estimation. Second, the analyses used summary statistics derived from the same cohort. Genetic correlation does not establish that cortical morphology causes externalizing behavior, that externalizing behavior alters cortical development, or that an imaging phenotype mediates genetic liability. Shared genetic effects could arise through pleiotropic or indirect developmental pathways. Third, regional imaging phenotypes are strongly correlated, making the effective number of independent comparisons smaller than the raw test count while also complicating interpretation of clusters of nominal results. Fourth, the findings have not yet been replicated in an independent developmental cohort. Finally, the ancestry composition of the GWAS and the LD reference panel must be clearly documented because LD Score Regression requires the LD reference to be appropriate for the population represented in the GWAS summary statistics.

Larger developmental imaging-genetics samples will be required to distinguish modest shared genetic effects from estimation noise. Future studies could improve precision by combining compatible imaging cohorts, examining hemisphere-specific measures, using multivariate imaging phenotypes, and testing whether promising regional results replicate in independent datasets. The nominal associations identified here, particularly those involving entorhinal and superior frontal cortical-thickness phenotypes, may be used to prioritize such replication analyses, but they should not currently be interpreted as established associations.

## Conclusion

In this initial analysis of the ABCD cohort, no individual genetic correlation between externalizing behavior and cortical imaging phenotypes remained significant after correction for multiple comparisons. The strongest nominal associations involved entorhinal, superior frontal, insular, and other cortical-thickness phenotypes, but these findings require independent replication.

Equivalence testing showed that only a small proportion of comparisons were sufficiently precise to support genetic correlations smaller than |*r*_*g*_| = 0.20. Most results were therefore inconclusive rather than evidence of no shared genetic influence. Overall, the findings indicate that substantially larger imaging-genetics samples and more precise SNP-heritability estimates will be needed to determine whether externalizing behavior and adolescent cortical development share modest or regionally specific common-variant genetic influences.

## Data availability

### Code

The analysis code and scripts used in this study are publicly available at: https://github.com/mw742/ext_ldsc_brain

### Data

This study uses data from the Adolescent Brain Cognitive Development (ABCD) Study (https://abcdstudy.org), available through the NIMH Data Archive (NDA). The ABCD data release used was version 5.1. The study is supported by the National Institutes of Health (NIH) and additional federal partners under multiple award numbers, including U01DA041048 and U01DA050987. A full list of funders is available at https://abcdstudy.org/federal-partners.html.

## Author contributions statement

Mengman Wei conceived the study, designed the analytical framework, performed data processing, statistical analyses, and computational modeling, and drafted the manuscript. Mengman Wei also carried out code implementation, data curation, and interpretation of the results. Qian Peng provided supervision, overall guidance, resource support, and funding acquisition.

## Preprint Notice

This manuscript is a preprint and has not yet undergone peer review. The content is shared to disseminate findings and establish a precedent. Additional analyses and revisions may be incorporated in future versions.

